# Fully Automated Delineation of the Optic Radiation for Surgical Planning using Clinically Accessible Sequences

**DOI:** 10.1101/2020.07.16.20155598

**Authors:** Lee B Reid, Eloy Martínez-Heras, Jose V Manjón, Rosalind L. Jeffree, Hamish Alexander, Julie Trinder, Elisabeth Solana, Sara Llufriu, Stephen Rose, Marita Prior, Jurgen Fripp

## Abstract

1

Quadrantanopia caused by inadvertent severing of Meyer’s Loop of the optic radiation is a well-recognised complication of temporal lobectomy for conditions such as epilepsy. Dissection studies indicate that the anterior extent of Meyer’s Loop varies considerably between individuals. Quantifying this for individual patients is thus an important step to improve the safety profile of temporal lobectomies. Previous attempts to delineate Meyer’s Loop using diffusion MRI tractography have had difficulty estimating its full anterior extent, required manual ROI placement, and/or relied on advanced diffusion sequences that cannot be acquired routinely in most clinics. Here we present CONSULT – a pipeline that can delineate the optic radiation from raw DICOM data in a completely automated way via a combination of robust preprocessing, segmentation, and alignment stages, plus simple improvements that bolster the efficiency and reliability of standard tractography. We tested CONSULT on 694 scans of predominantly healthy participants (538 unique brains), including both advanced acquisitions and simpler acquisitions that could be acquired in clinically acceptable timeframes. Delineations completed without error in 99.4% of the scans. The distance between Meyer’s Loop and the temporal pole closely matched both averages and ranges reported in dissection studies for all tested sequences. Median scan-rescan error of this distance was 1mm. When tested on two participants with considerable pathology, delineations were successful and realistic. Through this, we demonstrate not only how to identify Meyer’s Loop with clinically accessible sequences, but also that this can be achieved without fundamental changes to tractography algorithms or complex post-processing methods.

**Highlights:**

- We propose a fully automated means of delineating the optic radiation using diffusion MRI from DICOM data
- Anatomical measurements from tractography of over 500 brains align well with previous dissection studies
- The proposed pipeline works well with clinically accessible and advanced acquisitions
- Median scan-rescan error was 1mm
- Plausible tractography was generated when pathology was present

## 2 Introduction

The axons of the visual pathway emanate from the retina, partially bifurcate at the optic chiasm, pass on either side of the midbrain, and synapse in the lateral geniculate nucleus (LGN). Subsequent axons emanating from the LGN form the posterior, central and anterior bundles, referred to collectively as the optic radiation, which synapse in the primary visual cortex,^1^ and potentially V2 and V3.^2^ The posterior bundle travels almost directly posterior from the LGN, whilst the central bundle first travels more laterally, before following a similar path. The anterior bundle, which carries information from the superior visual quadrant, travels anteriorly, inferiorly, and laterally as it fans.^1^ It then curves around the lateral ventricle to travel posteriorly to the visual cortex. The entire optic radiation is critical to normal vision, but the anterior bend of the anterior bundle, often referred to as ‘Meyer’s Loop’, has received ongoing focus in neurosurgical science for two related reasons. Firstly, unlike other parts of the optic radiation, it is frequently severed during anterior temporal lobectomies and selective amygdalohippocampectomies, which are common treatments for epilepsy, resulting in permanent quadrantanopia.^3^ Such partial blindness can result in a reduced quality of life and independence, particularly by removing patients’ legal eligibility to drive.^3,4^ Secondly, radiologists can use standard structural images to localise vision-critical gyri in the occipital lobe and often even posterior aspects of the optic radiation itself (which appears as a band; Suppl Fig 1). By contrast, the majority of Meyer’s loop is not identifiable on such images.

The primarily investigated morphological measurement for the optic radiation has been the distance between the most anterior point of Meyer’s Loop and the temporal pole (ML-TP distance).^3^ This reflects the fact that temporal lobe resection commonly removes tissue from this zone, and this metric can inform the maximally appropriate posterior extent of tissue excision. Six influential cadaver dissection studies have suggested that this distance varies considerably between patients, ranging between 15 – 34mm.^1,5–9^ The reported average distances have been 25mm (n=20),^6^ 26mm (n=11),^8^ and 27mm (n=25),^5^ 28.4mm (n=20), and 31.4mm (n=10),^7^ giving a weighted mean of 27.2mm. It is important to remember that it is difficult to delineate the precise anatomical boundaries via the Klinger dissection technique used in these studies, meaning that some of these estimates could be slightly overestimated.^5^ Regardless, the range of distances reported implies that measurement of an individual surgical candidate’s ML-TP distance is more prudent than relying on population averages.

To achieve maximal clinical utility, a tool that delineates the optic radiation needs to be non-invasive, accurate, reliable, and not require costly resources such as manual input, training, or special hardware. Given its non-invasive nature, diffusion MRI tractography is an obvious option, and several groups have delineated the optic radiation using this technique.^2,10–22^ Notably, Chamberland et al.^10^ presented an imaging method that produced arguably the most realistic range of ML-TP distances to date. The success of this method, however, relied on manual ROI placement and a multishell sequence with b-values up to 5000 s/mm^2^, potentially impeding widespread clinical adoption. Such clinical-accessibility issues have proven difficult to overcome: to the authors’ knowledge, all published methods to date have required manual input (e.g. region-of-interest [ROI] placement and adjustment, skull stripping or other preprocessing) from trained technicians, and/or advanced MR sequences that rely on MRI hardware or multiband packages that are often unavailable clinically.

Beyond clinical accessibility, perhaps the largest challenge has proven to be achieving reasonable accuracy and reliability in optic radiation delineations. As noted by Chamberland, almost all published works reporting ML-TP distances, regardless as to their automaticity or acquisitions, have tended to at least somewhat underestimate the anterior extent of Meyer’s Loop (i.e. overestimate ML-TP distances).^10,21^ For example, Kammen et al^23^ described a sophisticated pipeline that automated all but image preprocessing, alignment, and skull-stripping steps. When applied to diffusion images that would take over an hour to acquire on a typical hospital scanner, this produced visibly convincing tractography but a mean ML-TP distance around 4mm greater than most dissection studies. Such difficulties have raised the question as to whether well-established tractogram generating algorithms are simply incapable of reliably delineating the optic radiation’s complex morphology. A number of groups have improved optic radiation delineations through fundamental changes in the way tractography is interpreted, performed, or filtered after generation.^16–18,21,23^ However, it is not settled as to whether such purpose-designed algorithms are fundamentally required for optic radiation delineation, or whether standard tractography could be capable of this task if confounding issues were resolved, such as being able to reliably delineate the typically invisible boundaries of the LGN.^17^

Herein, we describe an image processing pipeline central to our Connectivity Based Neurosurgical Planning Toolkit (CONSULT), which automatically processes images from their raw form and produces a binary map of the optic radiation suitable for surgical planning. For the clinical reasons discussed earlier, the focus of this pipeline, and its assessment here, is an accurate estimation of the ML-TP distance. CONSULT’s primary strength is that it is highly clinically accessible: it processes data from a range of sequences in their raw (DICOM) form, without requiring any human input, and produces ML-TP distances closely in line with historical dissection studies. We describe its stages and report ML-TP distances and reproducibility for a large sample of predominantly neurologically normal participants. Through this, we demonstrate that adequate estimates of ML-TP distance for surgical planning can be achieved without manual input or specialised sequences that require specialised scanner capabilities and impractical scan times.

## 3 Methods

We processed several datasets using the CONSULT pipeline, measuring the ML-TP distance in 538 unique brains. Within this pipeline are a small number of changes made to MRtrix3 that are designed to improve the efficiency of streamline generation but do not alter the underlying principles of the probabilistic tractography algorithm itself. We now describe the data processed, the changes made to MRtrix3, then the pipeline itself.

### 3.1 Data Processed

Table 1 details the scans processed, grouped into 6 datasets. The three largest datasets were participants from the Human Connectome Project (HCP),^24,25^ whilst three more were data acquired on scanners in clinical facilities. Approval was granted for data collection and acquisition from the relevant ethics committees in Metro North Australia, Hospital Cliníc de Barcelona, Washington University, and CSIRO. All participants gave written informed consent. Scan times listed below assume full-brain coverage, but approximately half this many slices is enough to image the optic radiation.

**Table 1.** Input image properties for the datasets used, ordered by number of diffusion directions. Changes were made to HCP data to produce the ‘HCP Multishell 60’ (HCP-M60) and ‘HCP Single Shell’ (HCP-S) datasets. ‘Preprocessed’ indicates that preprocessed images were used rather than raw images. Direction Repeats indicates whether any measurement repeats of each diffusion-weighted gradient direction were included in the data. Shell b-values are in s/mm^2^. ‘Acceptable TA’ indicates whether the sequence could be acquired in a clinically acceptable timeframe. The HCP Connectome Skyra is a modified Siemens Skyra scanner with differing performance to the commercially available Skyra model. †Acceptable TA using a Prisma or Vida scanner, or using multiband, or with partial brain coverage. Abbreviations: HCP, Human Connectome Project; HCP-M, Human Connectome Project multishell; ML-TP, Meyer’s-Loop to Temporal Pole distance; Preproc, minimally preprocessed data; Rev-P, reverse-phase encoding; TA, acquisition time.

| Name | Purpose | Scanner | Participant count | Time points | Coil Channels | Diffusion |  |  |  |  |  |  |  |  |  | T1 MPRAGE |
| --- | --- | --- | --- | --- | --- | --- | --- | --- | --- | --- | --- | --- | --- | --- | --- | --- |
|  |  |  |  |  |  | Data Used | Resolution | Unique Directions per Shell |  |  |  | Direction Repeats | Distortion Correction | Acceptable TA | Data Used | Resolution |
|  |  |  |  |  |  |  |  | b=0 | b=1000 | b=2000 | b=3000 |  |  |  |  |  |
| HCP-M90 | ML-TP against prior dissection studies in very high-quality data | Connectome Skyra | 500 | 1 | 32 | Preproc. | 1.25mm | 18 | 90 | 90 | 90 | Included | Rev-P | No | Preproc. | 1mm |
| HCP-M60 | ML-TP reproducibility in high quality data | Connectome Skyra | 40 | 2 | 32 | Raw | 1.25mm | 12 | 60 | 60 | 60 | Removed | Rev-P | No | Preproc. | 1mm |
| Hospital-A | ML-TP against prior dissection studies in high quality raw data | Siemens Prisma (Barcelona) | 19 | 1 | 64 | Raw | 1.5mm | 5 | 30 | 60 | 90 | Not Acquired | Fieldmap | With Prisma & Multiband | Raw | 0.9mm |
| Hospital-B | Qualitative assessment in presurgical patients collected in a clinically acceptable timeframe | Siemens Prisma (Brisbane) | 5 | 1 | 64 | Raw | 2mm | 12 | 20 | 32 | 60 | Not Acquired | Rev-P | Yes† | Raw | 1mm |
| Hospital-C | Demonstration using hardware and sequences without multiband | Siemens Biograph mMR (PET/MRI) | 2 | 1 | 8 | Raw | 2.5mm | 12 | 20 | 0 | 60 | Not Acquired | Rev-P | Yes | Raw | 1mm |
| HCP-S | ML-TP reproducibility and population norms in lower quality data | Connectome Skyra | 40 | 2 | 32 | Raw | 2mm | 2 | 0 | 0 | 60 | Removed | Rev-P | Yes | Preproc. | 1mm |

To assess the ML-TP distance in many participants, we processed the first 500 participants from the HCP Young Adults 1200 release (99% aged 22 – 35y, 40% male). This dataset is named within as ‘HCP Multishell-90’ (HCP-M90). Due to the large number of scans, preprocessed images were used due to reduce the computational cost to an achievable level.

To assess the reproducibility of CONSULT pipeline on high quality *raw* data, the ‘HCP Multishell 60 Repeat’ (HCP-M60) dataset was used. This consisted of 40 participants (28% male, aged 22 – 35y) from the Young Adults Release that were acquired at two time points. We processed both time points from raw diffusion data, but discarded all but 60 volumes per shell prior to preprocessing (see Table 1). Directions were removed by selecting sub-shells that each had an approximately evenly distributed set of directions using a freely available script (https://bitbucket.csiro.au/projects/CONSULT/repos/dwi_tools/). An equivalent sequence for a Siemens Prisma scanner equipped with multiband would carry an acquisition time of approximately 38 minutes.

The ‘HCP Single Shell’ (HCP-S) dataset was processed to assess the reproducibility and performance of the CONSULT pipeline with a lower quality dataset. This dataset consisted of the same participant data as the HCP-M60 dataset, but prior to preprocessing, we removed all but 2 × b=0 volumes and 60 × b=3000 s/mm^2^ volumes using the aforementioned script and downsampled this raw diffusion data to 2mm isotropic. An equivalent sequence can be acquired in ∼12 minutes on a Siemens Prisma scanner without multiband (∼6 minutes with multiband factor of 2).

We also sought to determine CONSULT pipeline’s performance on raw data acquired at clinical sites in clinically acceptable timeframes. The ‘Hospital-A’ dataset consisted of three-shell images from 19 neurologically healthy participants aged 21 – 45y (mean: 33.7y), and is regularly acquired at Hospital Clínic, Barcelona. Scan time for diffusion images and the associated field map was 20 minutes using multiband. The Hospital-B dataset consisted of three-shell images of five adult neurosurgical candidates (two with temporal lobe epilepsy, two with low grade gliomas, one with an arteriovenous malformation) acquired at the Royal Brisbane and Women’s Hospital campus, using a sequence that is regularly acquired at several Australian hospitals as part of clinical research (TA: 11.5 minutes with multiband, 23 minutes without). Results for Hospital-B were assessed qualitatively as genuine deviations from normal morphology could be expected in several cases. Finally, the Hospital-C dataset consisted of a low-resolution two-shell scan of two healthy adults acquired on a Siemens Biograph mMR scanner located on the Royal Brisbane and Women’s Hospital Campus (TA: ∼17 minutes without multiband).

### 3.2 Computationally Efficient Tractography

Seeding naively from near the LGN can convincingly generate the anterior bundle but only if left running for a very long time, as streamlines are far more likely to follow the central and posterior bundles, or aberrant paths, than successfully complete Meyer’s Loop. Although constraints may eliminate erroneous streamlines, tractography’s processing time remains prohibitive because such constraints rarely prevent most computational time being spent generating streamlines that are ultimately rejected, falsely giving the impression that delineating such complex anatomy is beyond the capabilities of existing tractography algorithms. We have forked MRtrix3^26^ and added three features that can improve efficiency but do not alter the nature of tractography itself. Source code for this forked version can be found at https://bitbucket.csiro.au/projects/CONSULT/repos/mrtrix3/. For brevity, detailed reasoning for these features can be found in Supplementary Materials.

Firstly, we propose a method by which seeding directions can be set in a voxel-wise manner from a unipolar dixel image (I.E. where each voxel contains a histogram of unipolar-direction probabilities) and converts pre-existing tractography to such images. This theoretically allows selective use of voxelwise seeding directions that are expected to be fruitful in generating a tractogram, including in a way previously demonstrated by mesh-based seeding.^27^

Secondly, we have added an option to tckgen that ensures that streamlines pass through inclusion regions in the correct order; this feature has also been pulled into the official distribution of MRtrix. Anatomically constrained tractography, an established method, can rewind streamlines when they terminate unsatisfactorily, allowing a re-attempt at completing.^28^ We have enabled ‘back-tracking’ within MRtrix for more general use by allowing this to run outside of anatomically constrained tractography, and requiring rewinding to explicitly defined ROIs that can be placed before problematic branch points. Here, we conservatively set the maximum number of back-track attempts at two per streamline.

### 3.3 Structural Processing

#### 3.3.1 Template Generation

Before processing, we created population probability images for labelling of the temporal lobe and V1/V2 region by affine registering 138 HCP ‘Young Adult’ participants to MNI space.^29^ Freesurfer-defined labels from these datasets were transformed into MNI space, and their mean taken to represent population probabilities for the temporal lobe and V1/V2 zones. Of these 138 scans, 54 participants also belonged to the HCP-M90 dataset. No participants within HCP-M60, HCP-S, or Hospital datasets contributed to such atlas creation.

#### 3.3.2 Basic Processing

For all Hospital datasets, raw T1 scans were N4 bias corrected^30^ and skull stripped using HD-BET.^31^ Global Approximate Block Matching was used to denoise and reduce any motion artefacts, as well as segment these images into grey matter, white matter, and cerebrospinal fluid.^32,33^ A non-linear transform from MNI space was also calculated using ANTs SyN^34^ using a multi-atlas method.

Due to the unusual bias-correction requirements for HCP structural data,^25^ minimally preprocessed T1 data were used for all HCP datasets. To reduce computational overhead these scans were resampled to 1mm isotropic. These data were then processed identically to the hospital datasets, excepting that bias-correction was not (re-)performed. No masks, registrations, or parcellations provided with the HCP data were used for either the structural or diffusion data, except for generation of population templates (see above).

### 3.4 Diffusion Processing

#### 3.4.1 Preprocessing

The HCP-M90 dataset was used in its minimally preprocessed form to ensure an achievable computational cost given the large numbers of scans processed. This preprocessing included correction for b0 intensity inhomogeneities, EPI distortion, eddy currents, head motion, gradient non-linearities, as well as reorientation and resampling to 1.25mm isotropic.^25^

Data from all datasets except HCP-M90 were preprocessed by CONSULT from their raw form as follows. Images were denoised using MRtrix’s dwidenoise, and motion-affected volumes were removed using a previously described method.^35^ If a fieldmap was detected (here, only the Hospital-A dataset), a brainmask was calculated from the magnitude phase image using bet2^36^ as this is more robust to signal dropout in the temporal lobe, and a fieldmap calculated using fsl_prepare_fieldmap. Alternatively, for reverse-phase encoded scans (all but the Hospital-A dataset), an initial brainmask was calculated using MRTrix’s dwi2mask, but dilated to ensure full temporal-lobe coverage, and spatial distortions were calculated using FSL’s topup (http://fsl.fmrib.ox.ac.uk/fsl/fslwiki/TOPUP). For all datasets motion, distortion, and eddy-current correction were performed through eddy_cuda 8.0. Intensity inhomogeneities were also corrected using N4 bias correction, the field being estimated from the eddy-corrected mean b0 image. For reverse-phase encoded scans, a final brainmask was then recalculated using dwi2mask and dilation operations.

#### 3.4.2 Processing

After preprocessing, all datasets were processed identically, unless otherwise stated. Tissue response functions were calculated using the Dhollander algorithm.^37^ Fibre orientation dispersion maps were calculated for white matter, grey matter, and cerebrospinal fluid using multishell multitissue contrained spherical deconvolution (multishell data) or Single-Shell 3-Tissue constrained spherical deconvolution (HCP-S dataset; https://3Tissue.github.io). Fractional anisotropy was calculated using MRtrix3, upsampled by a factor of 2 using sync interpolation, and cropped to the final brainmask.

Several ROIs, defined in MNI space, were needed for tractography (see below). To transform these into native diffusion space, a transform from diffusion to MNI space was calculated: the fractional anisotropy image was rigid registered to the participant’s T1 image using ANTs and combined with the previously-calculated non-rigid transform from T1 space to MNI space.

### 3.5 Posterior Bundle

To aid identification of the anterior bundle, the posterior bundle, and a partial central bundle, were delineated by tracking unidirectionally from the optic tract to V1/V2. Streamlines were generated until Tractogram Bootstrapping^38^ stability criteria were met (min Dice, 0.95; reliability, 0.95; resolution equal to diffusion spatial resolution; b_int_, 0.001×n; minimum streamline count, 10000). The V1/V2 region was calculated by transforming the previously described V1/V2 template from MNI into native diffusion space, multiplying by individual participant’s grey-matter probability image taken from the FOD maps, thresholding and binarising. Exclusion masks included the grey-matter segmentation (excepting in primary visual areas) and a mask defined in MNI space similar to that described by Martinez-Heras et al.^15^

We seeded from the optic tract near the midbrain, which was identified using a convolutional neural network, as we found seeding from the optic chiasm^23^ was sometimes impaired by MR artefacts. Details of this network are supplied in Supplementary Materials. In brief, the T1 and white matter fibre orientation dispersion images were transformed into MNI space and cropped to a stack of six 1mm-thick axial patches in the centre of the brain. These were provided to this network, which had a UNet-style architecture^39^ and had been trained with datasets for which the optic tract had been identified using tractography seeded from the optic chiasm. The output was a binary mask of the optic tract as it entered the cerebral hemispheres, which could then be padded and transformed back into native participant space (Suppl Fig 3).

### 3.6 Central and Anterior Bundles

The remaining partial-central and anterior bundles were delineated by tracking from the LGN to the visual cortex. This was performed in a two-pass procedure. In both instances, exclusion ROIs were those previously described.

#### 3.6.1 First Pass: Identifying an Efficient Seed Point

The first pass was designed solely to identify a seeding position and direction near the LGN, for efficient delineation of the anterior bundle during the second pass. A seeding region was calculated by binarising tractography of the posterior bundle (Figure 1A-B), multiplying by an ROI slightly anterior to the LGN (transformed from MNI space; Figure 2, top) and dilating once (Figure 1C). This mask was slightly anterior to the true LGN because directly seeding from the LGN can meaningfully slow streamline generation. Tractography was then seeded from this ROI (Figure 1D), the streamline count being set automatically by Tractogram Bootstrapping^38^ for a reproducible map of streamline directions (I.E. unipolar-dixel trackmap; default settings used and minimum streamline count of 1000). Three ordered inclusion regions were used: the seed ROI, the temporal lobe posterior and medial to the expected tip of the anterior bundle (Figure 2, middle), and a pre-occipital plane (Figure 2, bottom). To improve efficiency, the second inclusion region was used as a back-tracking ROI. This back-tracking ROI allowed streamlines that entered the second inclusion region but were ultimately rejected to be rewound to approximately 44mm posterior to the temporal lobe and reattempt completion (up to twice per streamline).

**Figure 1.**
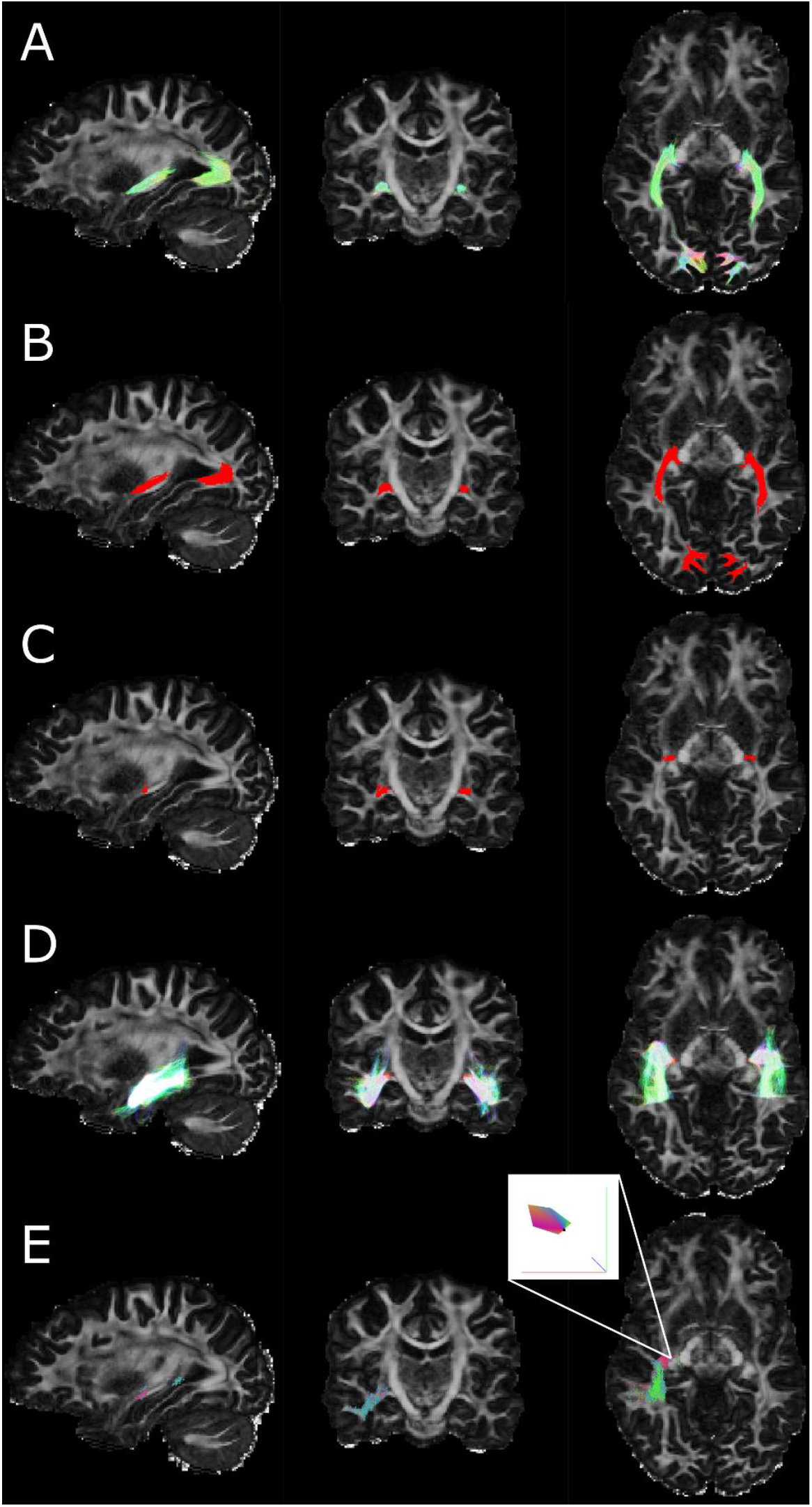
Steps related to the first pass generation of streamlines describing anterior and middle bundles. Tractography of the posterior bundle (A) was converted into a binary ROI (B), then masked to an ROI slightly anterior to the LGN (C). Tractography was then performed (D) and converted into a map of streamline directions (E). The inset in E shows a zoomed view of a single voxel containing a histogram of streamline directions (unipolar dixel). This dixel map was used to improve efficiency of the seeding operation for the second pass of tractography (see Results for examples). Note that tractography is displayed cropped to the shown slices.

**Figure 2.**
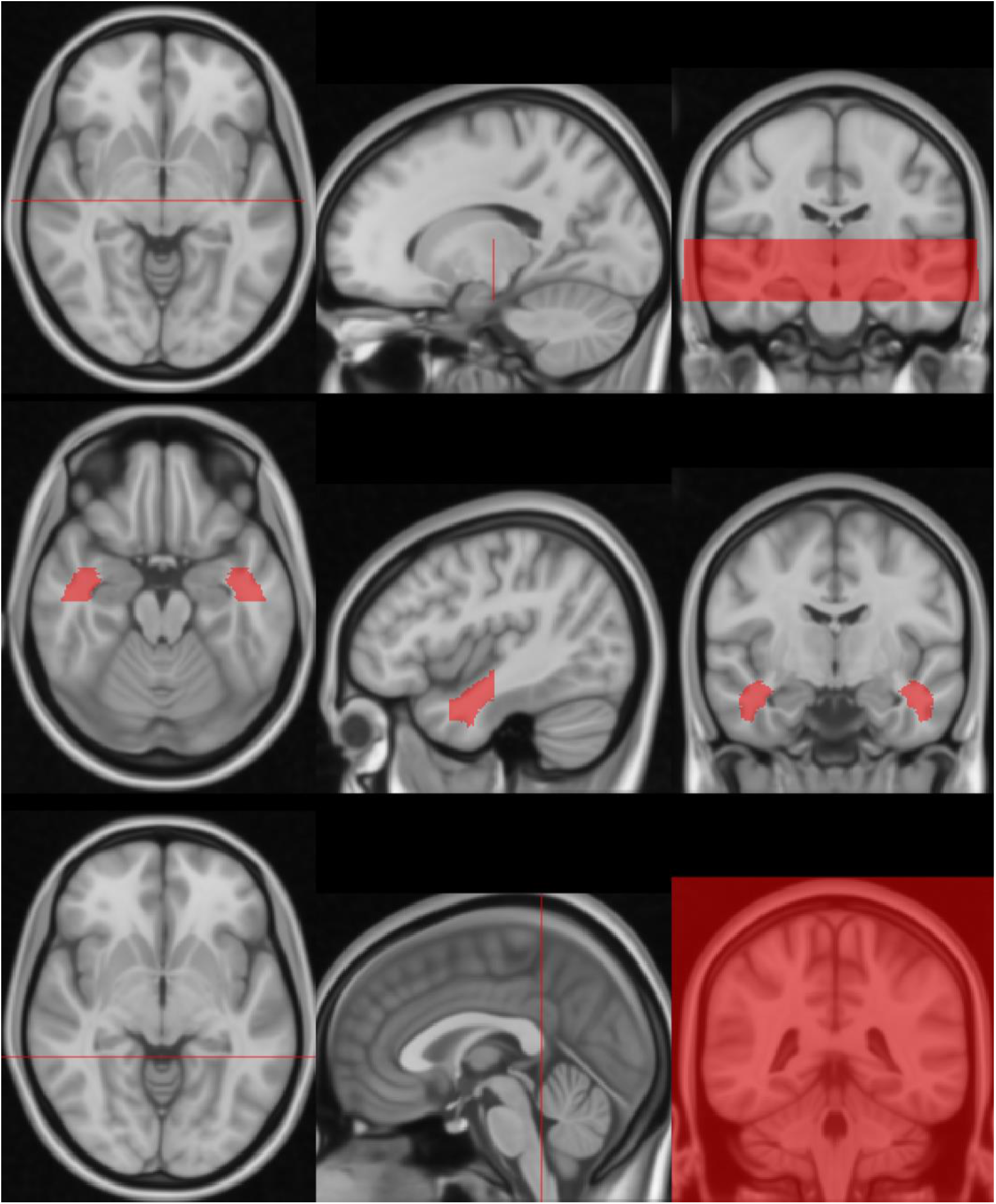
Ordered inclusion regions for the anterior bundle, as defined on the non-linear MNI152 template. Streamlines must pass through these in the order displayed here (from top to bottom). Top: the approximate pre-LGN plane, also used for generating the seed. Middle: the temporal lobe inclusion and backtrack region. Bottom: the pre-occipital plane. The fourth inclusion region was the visual cortex, which was generated more dynamically by combining a population average with the participant’s tissue segmentation (see text), and so is not displayed here.

#### 3.6.2 Second Pass: Tracking the Full Bundle

The tractogram generated by the first pass was converted into a unipolar dixel image in which each voxel represented a histogram of seeding directions that were successful in delineating Meyer’s loop (Figure 1E). The first-pass tractogram was then discarded. Streamline generation was seeded from the same ROI as the first pass, but with initial seeding directions sampled from this dixel map. Four ordered inclusion ROIs were used: the same three as the first pass, followed by the visual cortex ROI previously described. Streamlines were generated until Tractogram Bootstrapping^38^ stability criteria were met for a binary track map (min Dice, 0.95; reliability, 0.95; spatial resolution double that of the diffusion MR; b_int_, 0.001×n; minimum streamline count, 10000). The final tractogram was converted into a track density image, thresholded at 0.001 × number of streamlines generated and binarised, in line with the Tractogram Bootstrapping criteria.

### 3.7 Morphological Measurement

To enable fair comparisons between datasets in terms of brain volume, binarised tractograms were warped into MNI space^29^ for morphological measurements. We defined the ‘normalised ML-TP distance’ (nML-TP) as the anterior distance between the most anterior voxel of the temporal lobe (y = 25mm) and the most anterior voxel of the anterior bundle, in MNI space.

## 4 Results

### 4.1 Success Rates and Normative Statistics from the HCP

Typical per-scan processing times for 2mm multishell datasets were 3 – 4 hours on a 20 × 2.6Ghz core GPU-enabled machine, the majority of which was spent on diffusion preprocessing and registration steps. As data were processed on a network drive better performance can be expected on a dedicated machine. Data processing failed in 3 of 694 datasets (Table 2). Two failures were in HCP-M90 datasets: one due to a failed registration between the T1 and fractional anisotropy image, and another due to failed skull stripping in a participant displaying presumed cutis verticis gyrata. The third failure was an HCP-S dataset for which data processing completed but the nML-TP distance for one hemisphere was implausible (39 mm); this dataset was still included in all statistical analyses. Qualitatively, tractography aligned well with expected anatomy. The middle and posterior sections of the optic radiation aligned well with the corresponding band visible on T1 images, though its likely lateral extent was often slightly overestimated for acquisitions with spatial resolution > 1.5 mm (Suppl Fig 4). Normalised ML-TP distances are summarised in Table 2. The HCP-M90 dataset showed a median of 27mm and range (14mm – 33mm) in line with historical dissection studies.^7^ Despite this wide range, the interquartile range was tight (25 – 28mm) and 91% of measurements were within 4mm of the median. Median nML-TP distances did not differ between men and women.

**Table 2.** Normalised Meyer’s Loop to Temporal Pole Distances (nML-TP) and Data Failure Rates For Each Dataset. The 50% percentile refers to the median value. ‘Both’ hemispheres indicates that both hemispheres’ statistics have been pooled. HCP-M60 and HCP-S datasets show values for the first timepoint only. Spatial resolution is displayed in mm and volume in mL.

| Dataset | Hemisphere | Resolution | No<br>Datasets<br>Processed | Processing Failed | nML-TP (mm) Percentile |  |  |  |  |  |  | Median Anterior +<br>Central Bundle<br>Volume (mL) |
| --- | --- | --- | --- | --- | --- | --- | --- | --- | --- | --- | --- | --- |
|  |  |  |  |  | Min | 5% | 25% | 50% | 75% | 95% | Max |  |
| HCP-M90 | Both | 1.25 | 500 | 3 (0.6%) | 14.0 | 21.0 | 25.0 | 27.0 | 28.0 | 30.0 | 33.0 | 21.0 |
| HCP-M60 | Both | 1.25 | 40 | 0 (0%) | 19.0 | 21.0 | 25.0 | 26.0 | 28.0 | 29.0 | 31.0 | 20.0 |
| Hospital-A | Both | 1.5 | 19 | 0 (0%) | 24.0 | 25.0 | 27.0 | 28.0 | 29.0 | 30.0 | 30.0 | 24.1 |
| Hospital-B | Ipsilesional | 2 | 5 | 0 (0%) | 22.0 | 22.6 | 25.0 | 27.0 | 28.0 | 29.6 | 30.0 | 30.0 |
|  | Contralesional | 2 | 5 | 0 (0%) | 25.0 | 25.2 | 26.0 | 27.0 | 28.0 | 28.8 | 29.0 | 30.2 |
| Hospital-C | Both | 2.5 | 2 | 0 (0%) | 26.0 | N/A | 26.8 | 27.5 | 28.5 | N/A | 30.0 | 32.7 |
| HCP-S | Both | 2 | 40 | 1 (2.3%) | 20.0 | 23.0 | 26.0 | 28.0 | 29.0 | 31.0 | 39.0 | 25.0 |

Meyer’s loop typically extended marginally more anterior on the left side than the right (median nML-TP distance difference 1mm; p<0.001, one-sample Wilcoxon signed-rank test; Figure 6, middle). Such asymmetry did not differ in left-handed versus right-handed participants (Edinburgh Handedness Inventory score, < −0.2 vs > 0.2; medians both 1mm).

### 4.2 Reproducibility and Dataset Comparison

Healthy datasets showed visibly similar results to one another (Figure 3, Figure 4), as well as numerically similar medians and interquartile ranges for nML-TP distance (Table 2; Figure 6, top). When restricted to identical participants (n=28), median nML-TP distances matched for HCP-M60 and HCP-M90 (both 26mm; p = 0.48, Wilcoxon Signed Rank Test), but HCP-S showed a greater median nML-TP distance than HCP-M90 (28 vs 26mm; p<0.001, Wilcoxon Signed Rank statistic). Hospital-A’s median nML-TP distance was marginally greater than that of HCP-M90 (28mm vs 27mm; p<0.001, Mann-Whitney U Test). For the two Hospital-B scans showing gliomas, CONSULT was able to delineate plausible axonal pathways affected by nearby oedema (Figure 5) and produce nML-TP distances within the normal range for the contralesional hemisphere. Similarly, delineations were numerically and visually normal for both hemispheres of the remaining Hospital-B patients, and two healthy participants in Hospital-C. Despite producing similar anterior extents, lower resolution sequences typically resulted in physically-broader delineations than higher resolution sequences (Table 2).

**Figure 3.**
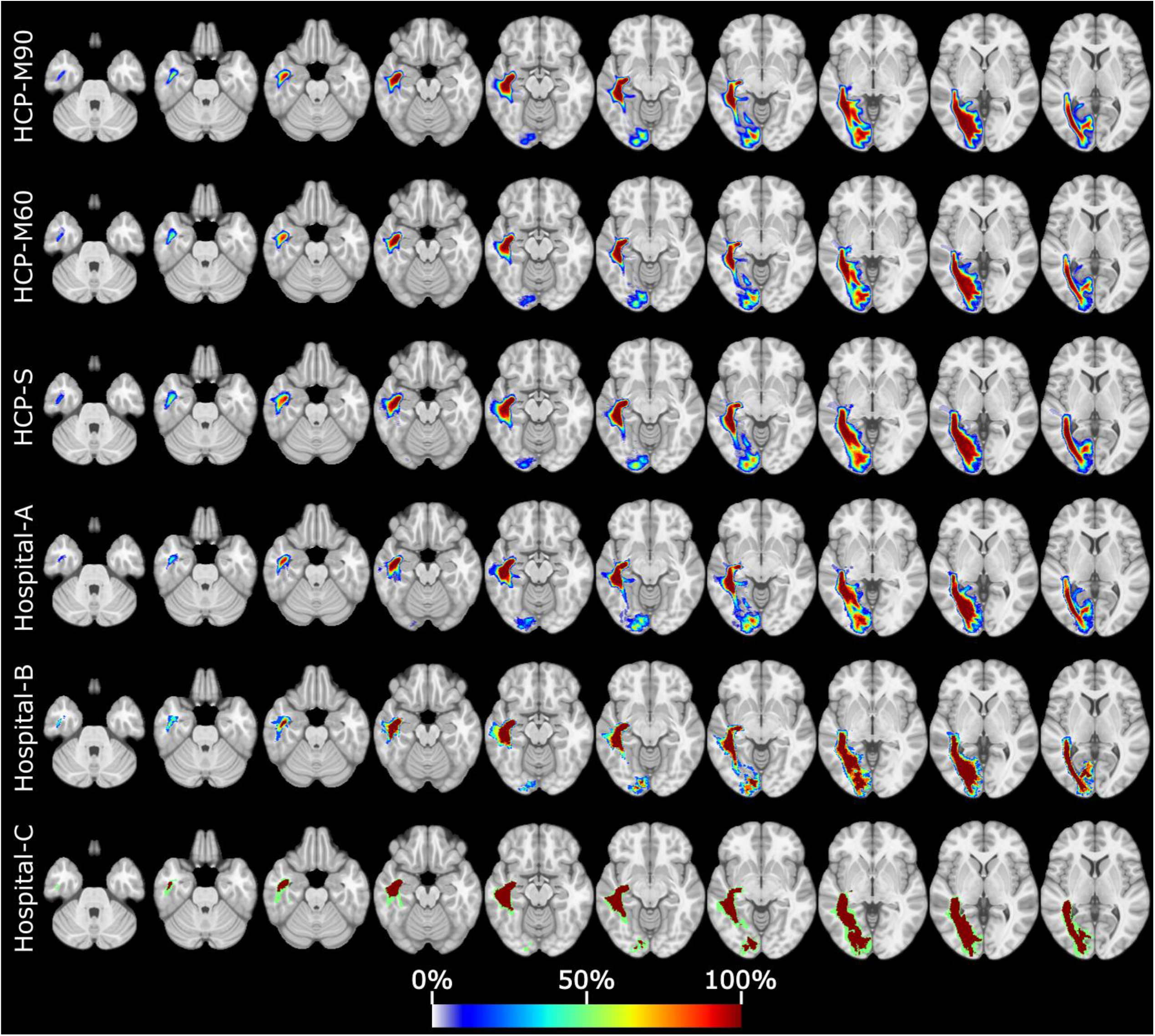
The right anterior bundle for all participants, pooled by dataset into rows, warped into MNI-152 space. Colour indicates the percentage of participants with binarised tractography in that location. Mean morphology is largely the same between all datasets. Hospital-C’s simple colouration is because this dataset only comprised two participants.

**Figure 4.**
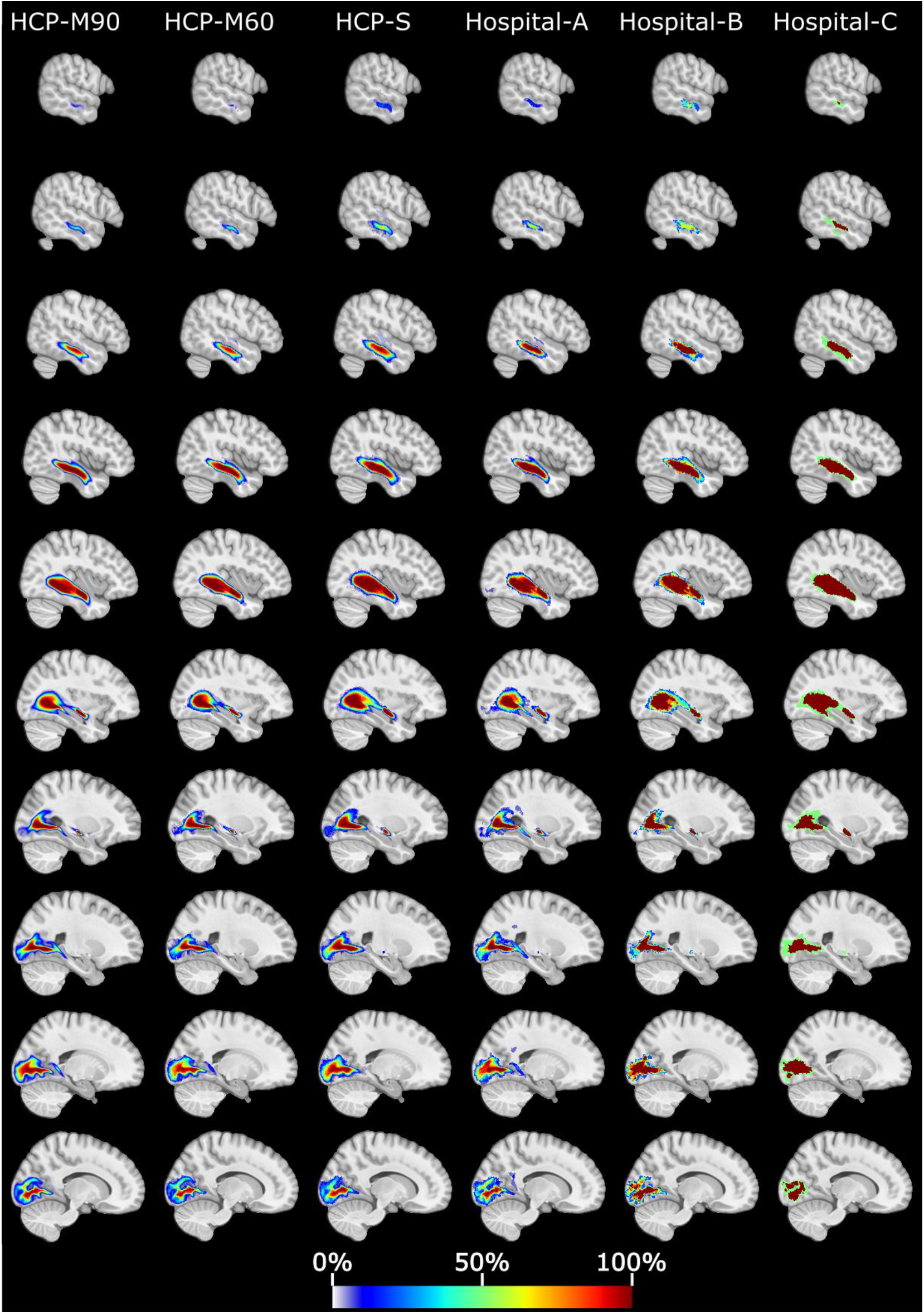
The right anterior bundle for all participants, pooled by dataset into columns, warped into MNI-152 space. Colour indicates the percentage of participants with binarised tractography in that location. Mean morphology is largely the same between all datasets. Hospital-C’s simple colouration is because this dataset only comprised two participants.

**Figure 5.**
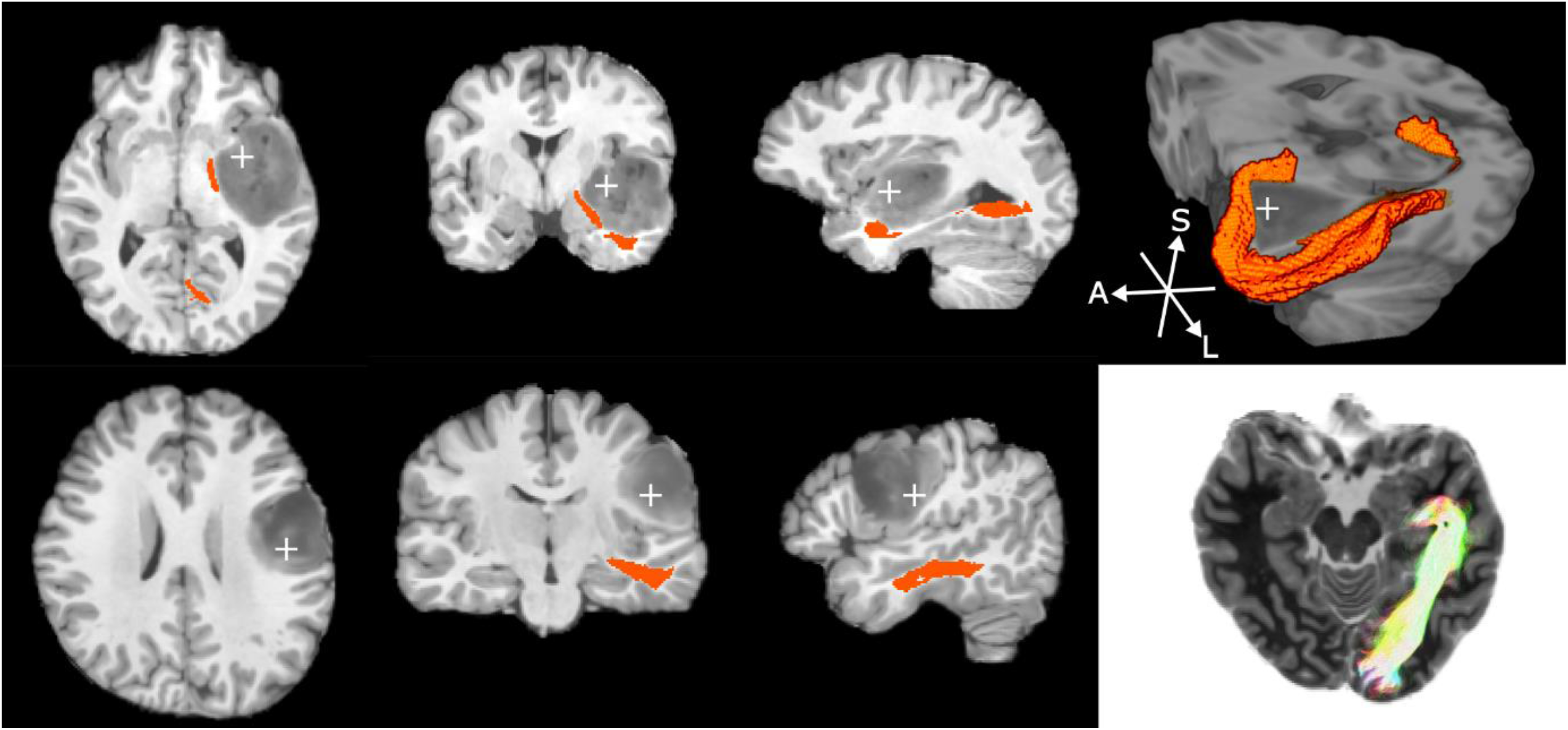
Pathology for two participants with gliomas in the Hospital-B dataset, shown on T1 MPRAGE scans. Colour indicates the delineated anterior bundle on the ipsilesional side. Top row: a neurosurgical candidate with pathology in the temporal lobe. The 3D cut-away image (right) shows the delineated Meyer’s Loop circling the region of oedema. Bottom row: a neurosurgical candidate with pathology to the frontal lobe causing some geometric distortion of the temporal lobe. The far-right image on this row is an axial slice with streamlines from all slices visible, and image intensities inverted to ease visualisation.

**Figure 6.**
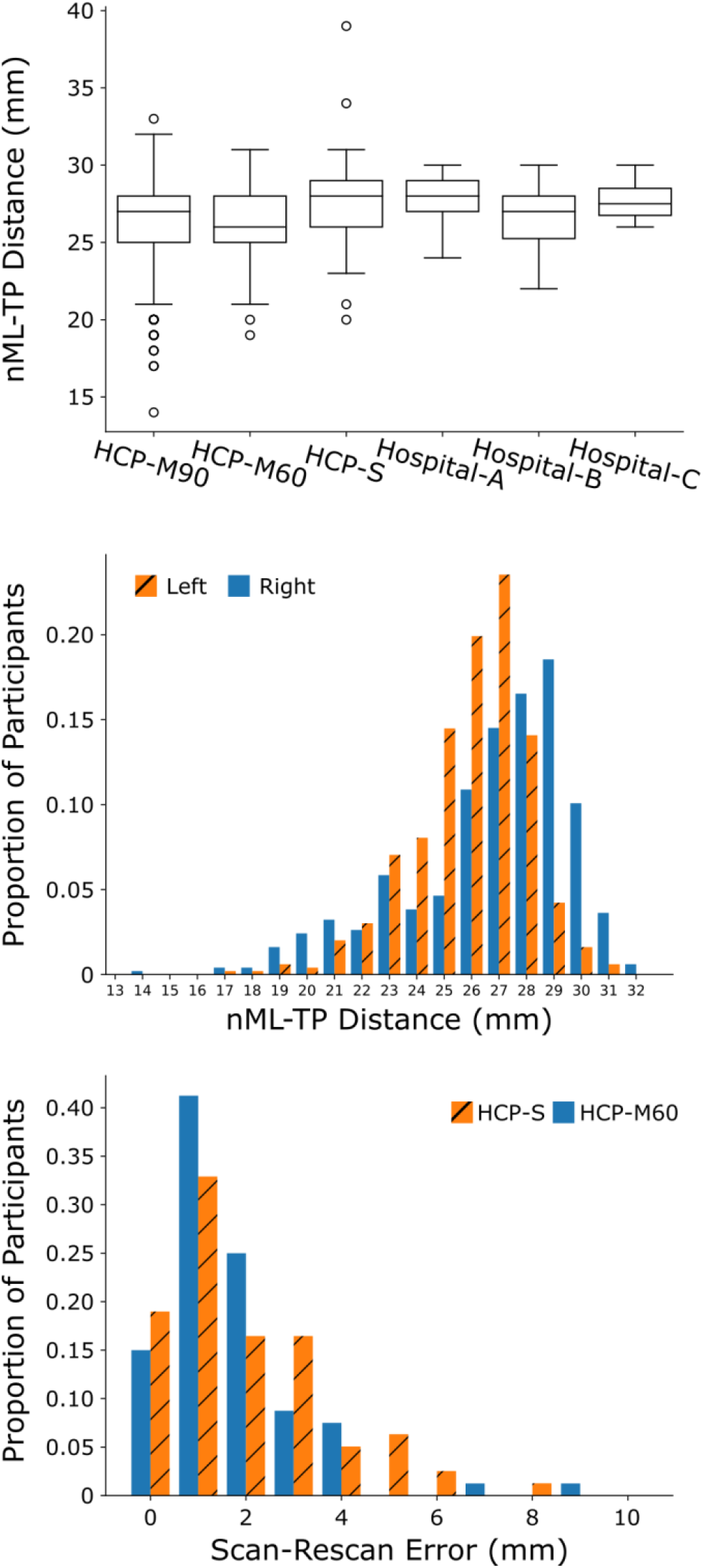
Top: Normalised Meyer’s Loop to temporal pole (nML-TP) distances for all processed datasets. Measurements are pooled from both hemispheres. Sequence details can be found in Table 1. Middle: nML-TP distances for the left and right hemispheres from the HCP-M90 dataset. Bottom: Absolute Scan-Rescan error for the HCP-S and HCP-M60. Note that more than 50% of scans showed error below the resolution of the raw images (2mm for HCP-S, 1.25mm for HCP-M60).

For the HCP-S and HCP-M60 datasets, we processed both scans of each participant that were available. Median absolute scan-rescan error in nML-TP measurements were 1mm for both datasets (Figure 6, bottom; p=0.15 Wilcoxon Signed Rank Test).

## 5 Discussion

We have demonstrated a pipeline primarily designed to delineate Meyer’s Loop, which is often inadvertently severed during brain surgery. CONSULT distinguishes itself from previously published works particularly in its ability to process raw data in a fully automated way. Here, CONSULT achieved nML-TP distances for a range of acquisitions (medians of 26 – 28mm) very similar to average distances reported in four of five dissections studies reporting this distance (25 – 28.4mm)^5,6,8,9^ and similar range (14 – 33mm, plus one failure at 39mm) to all five of such dissection studies (15 – 34mm).^5–8^ By comparison, most previously described MRI pipelines have reported average ML-TP distances larger than those reported in most dissection studies,^10^ implying that their clinical use may underestimate the risk of partial blindness associated with temporal lobectomy. For HCP-M60 and HCP-S, median scan-rescan of reproducibility of nML-TP distances was 1mm, though in ∼25% of cases was greater than 2mm, implying that it is prudent to artificially enlarge these delineations before providing them for clinical use. Our informal testing (not presented here) has suggested that the non-deterministic registration steps account for much of this variability, suggesting alternative registration algorithms may improve CONSULT’s reproducibility.

### 5.1 Clinical Accessibility

To achieve maximal clinical utility, an imaging tool that delineates the optic radiation needs to minimise its dependency on manual input, training, and special sequences/hardware. Manual input can be a high barrier to clinical adoption as it is time consuming, requires training, and interrupts existing clinical workflows. CONSULT is not the first pipeline published with automated components; most recently-described pipelines contain some degree of automation. For example, Kammen et al^23^ and Clatworthy et al^14^ both automated ROI placement, but also both required that supplied diffusion and structural images were already preprocessed, skull-stripped, and aligned to one another: steps that often require manual adjustment and quality control to ensure adequate performance. Other works have presumably automated several of these steps, but still required manual ROI placement. By contrast, CONSULT can process raw images without need for human input at any stage of the process and, as discussed below, has demonstrated an ability to process data acquired with a range of parameters and hardware.

Presentation is also important: visualising streamlines or streamline density images requires meaningful experience because streamline density does not strictly represent tissue density.^14^ As discussed in a previous publication,^38^ CONSULT’s conversion of streamlines into a binary map for visualisation may be beneficial for clinical decision making because it allows the boundaries to be defined mathematically rather than intuitively. This approach also allows results to be converted into DICOM format for viewing in standard clinical PACS-enabled viewers, minimising disruption to existing clinical workflows. Commendably, some earlier works converted tractography into binary maps,^14,15^ though tractography bootstrapping (applied here) carries the advantage of producing maps with better-quantified reproducibility.^38^

### 5.2 Simple versus Advanced Sequences

Some previous works have suggested that high b-value (b=5000 s/mm^2^) multishell sequences may be required to achieve a reasonable delineation of the optic radiation,^10^ and even if simpler acquisitions were sufficient, that there is no implication that standard tracking algorithms know how to propagate through the crossing regions of the optic radiation.^21^ Similarly, others have suggested that reasonable delineations require specialised track-filtering algorithms or thresholding techniques tuned to specific sequence/hardware combinations.^14,17,18^ If true, such suggestions would add significant barriers to achieving reliable delineations for most hospitals. Although probably beneficial, we found no evidence that such advances are absolutely required. Specifically, beyond a few efficiency boosting techniques, we were able to achieve plausible delineations using standard tractography algorithms, filtered by simply thresholding the resulting streamline density image without sequence-specific parameter tuning.^38^ More importantly, although we did not have an acquisition with a b=5000 s/mm^2^ shell for a direct comparison, we were able to produce such delineations using a variety of acquisitions collected on four scanners, including readily-accessible single-shell images. We recommend that multishell, or higher spatial resolution, images are used where possible, partially because of the slight differences observed between our HCP-M90 and HCP-S results and partially because a higher degree of directional redundancy from longer acquisitions affords some protection against motion artefacts. However, high resolution multishell images are clearly not a *requirement* because the anterior extent of Meyer’s loop, a variable but clinically-important measure for planning temporal lobectomies, differed only slightly between all of the tested acquisitions (Figure 6), and the reproducibility of this measure did not significantly differ between lower-resolution single-shell and high-resolution multishell acquisitions. Of course, most data here was acquired on scanners with good diffusion-gradient capabilities but, as stated in Section 3, several of these sequences can still be acquired on scanners with lower capabilities in clinically reasonable time frames. It is noteworthy that the Hospital-C scans were acquired on a scanner with poorer MR capabilities than most clinical scanners.

Despite estimating similar anterior extents, the lower resolution sequences produced broader delineations of the anterior loop than the lower resolution images despite tractography. This can be expected as result of more limited boundary information. Of course, in a surgical context where safety is paramount, labelling the entire Meyer’s Loop is more important than eliminating false-positive streamlines and so a mildly higher volume for these low-resolution sequences should not be considered a major drawback.

### 5.3 Limitations

One potential criticism of our work is that, like many others, we focussed on achieving a reasonable and reliable delineation of Meyer’s loop, but focussed less on quantitatively validating the middle and posterior sections of the optic radiation. This focus was because our motivation was surgery-specific: unlike the optic radiation’s posterior and middle sections, Meyer’s loop is more typically at risk from common neurosurgical operations that typically remove large amounts of the middle temporal lobe. The appropriate posterior extent of these incisions can be directly informed by the ML-TP distance, which requires specialised imaging techniques such as diffusion MRI. By contrast, as detailed in the Introduction, the medial and posterior sections of the optic radiation can be somewhat identified by more rudimentary means. Furthermore, demonstrating plausible Meyer’s Loop delineations in predominantly healthy individuals is a reasonable validation for planning temporal lobectomies because patients with epilepsy are the typical recipients of such treatments and characteristically have relatively normal appearing brains. Demonstrations in healthy controls for the posterior optic radiation, however, does not imply clinical success given that surgery near these areas is more likely to be for malformations that would cause a pipeline to fail.

A related limitation is that, given our datasets, it is not possible to validate our delineations against ground-truth data such as post-surgical outcomes. Validation in this sense is important for a clinical tool and ongoing work for our group, though some assurances should come from the average and range of morphological measurements found here closely matching several dissection studies, a challenge that the imaging community has only achieved a small number of times.^10,21^

### 5.4 Conclusion

We have presented a clinically accessible, fully automated pipeline for delineation of the optic radiation. This pipeline plausibly estimated the anterior extents of Meyer’s Loop in a range of hardware + sequence combinations, including both multishell and single-shell images that can be acquired in clinically acceptable time frames. We also demonstrated that, excepting some improvements in efficiency, such delineations can be achieved without resorting to complex streamline-selection criteria or fundamental changes to the way in which tractography is performed.

## Data Availability

Population averages can be requested from the authors. Due to the risk of de-identification, data from individual patients cannot be requested

## 5.5 Acknowledgements

This research was partially supported by an Advance Queensland Research Fellowship (R-09964-01), as well as the Spanish DPI2017-87743-R grant from the Ministerio de Economia, Industria y Competitividad of Spain. The authors gratefully acknowledge the support of NVIDIA Corporation with their donation of the TITAN X GPU used in this research, as well as Irene Pulido, Magí Andorrà Inglés, and other staff involved with subject enrolment and acquisition at HIRF and Hospital Clinic. Data were provided in part by the Human Connectome Project, WU-Minn Consortium (Principal Investigators: David Van Essen and Kamil Ugurbil; 1U54MH091657) funded by the 16 NIH Institutes and Centers that support the NIH Blueprint for Neuroscience Research; and by the McDonnell Center for Systems Neuroscience at Washington University.

## 7 Supplementary Materials

### 7.1 Discussion on novel MRtrix Features

#### 7.1.1 Voxel-wise Seeding Direction

Region-to-region tractography usually is executed by seeding from one ROI and keeping streamlines that arrive in a second ROI, which results in two forms of inefficiency. Firstly, with unidirectional seeding, at least half of all streamlines will travel the wrong direction from the seed. Bi-directional seeding does not prevent this but rather enforces it, causing streamlines to no longer begin in the desired ROI. Secondly, when seeding from a region containing more than one fibre population, streamlines may follow completely unrelated anatomy and so either become rejected, or by chance find the end region and be erroneously included in the final tractogram. Although the current implementation of MRtrix3 allows an initial tracking direction to be set, this is a single vector that is identically applied to all voxels, and so is not suitable for seeding from a curved surface (e.g. a gyrus) or from a region of fanning fibres (e.g. the LGN).

By seeding from a unipolar dixel image (I.e. where each voxel contains a histogram of unipolar-direction probabilities) time is not spent seeding in directions that are unlikely to produce rejected streamlines. This also affords the advantages previously provided by surface (mesh) based seeding.^27^

#### 7.1.2 Ordered Include Regions

Tractography between ROIs often utilises ‘include’ regions which must all be passed through for a streamline to be retained. Although very useful, the order these are passed through is not enforced, which can lead to erroneous inclusion of some streamlines. For example, when delineating the corticospinal tract, it is common for streamlines to leave the cortex, cross hemispheres, contact the brainstem, and travel superiorly to the original side’s internal capsule mask. These issues are often solvable with additional exclude regions, but their placement adds time and effort, and suitable locations often require trial and error, particularly in participants with pathology. For delineating the optic radiation, it is also common for streamlines to pass from the LGN to the temporal pole, then return to the LGN before passing to the primary visual cortex via the posterior bundle. We have added “ordered include” regions to the current options in tckgen to circumvent these issues.

#### 7.1.3 Explicit Backtracking

In regions where fibres branch, it is common for streamlines to follow the wrong anatomy and ultimately be excluded from analyses. This inevitable consequence of probabilistic tractography is wasteful in that the section before the branch may be accurate but this poor branching behaviour sees the entire streamline rejected. Anatomically constrained tractography, an established method within MRtrix, can rewind streamlines when they terminate unsatisfactorily, allowing a re-attempt at completing. We have enabled ‘back-tracking’ within MRtrix for more general use by allowing this to run outside of anatomically constrained tractography, and requiring rewinding to explicitly defined inclusion ROIs that can be placed before problematic branch points. In the present study the maximum number of back-track attempts per streamline was conservatively set at two.

We have informally tested CONSULT without use of back-tracking. Without this feature, tractography typically produces virtually identical delineations, but in some instances can take 2 – 3 times as long to complete. This is particularly prevalent when pathology is present.

### 7.2 The Optic Radiation as Visible on Structural Images

**Suppl Fig 1.**
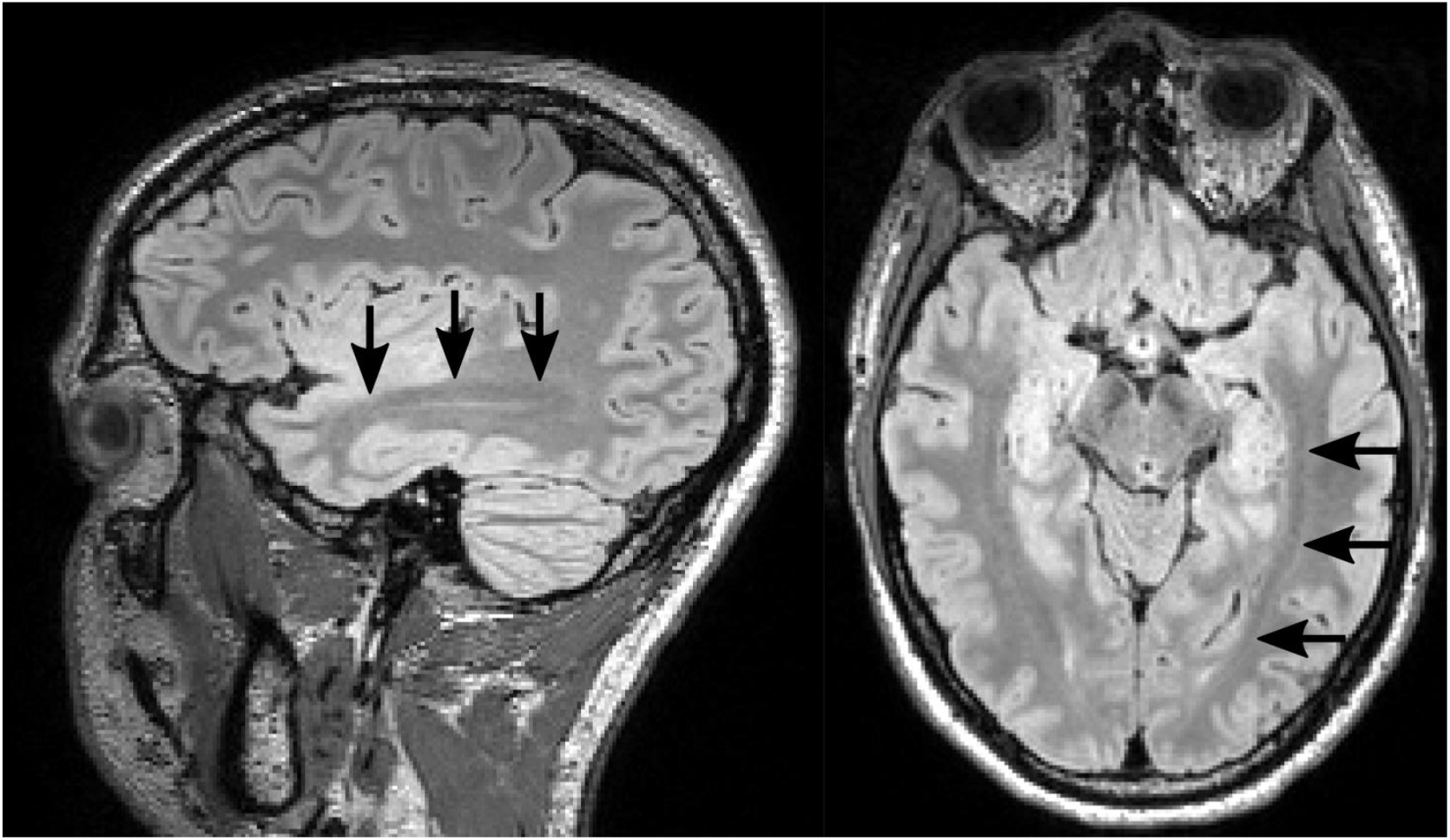
A raw FLAIR image of a neurosurgical patient. Much of the middle and posterior optic radiation (arrows) is visible as a band.

### 7.3 Optic Nerve Identifying Convolutional Neural Network

The optic chiasm is typically visible on fractional anisotropy (FA) images, providing a potential seeding point for tractography to identify the LGN or posterior bundle of the optic radiation. In unpublished analyses, we have previously identified a seeding point by transforming an approximate optic-chiasm region from MNI space, multiplying this by the upsampled FA image, thresholding at 0.3, and binarizing the result. In our experience, this method works on a variety of datasets, but skull stripping at times can remove the optic chiasm and the Hospital-A dataset presented here contained an artefact localised to this region in several instances, limiting the utility of this method. To overcome this, for all datasets in this study, we instead seeded from the optic tract near the brainstem.

To identify the optic tract, we trained a convolutional neural network. This network accepted T1 and white matter fibre orientation dispersion images from a single participant, after a preprocessing stage affine registered these to MNI space (1mm isotropic) and cropped them to six axial slices in the centre of the brain. The output was a binary mask of the optic tract as it entered both cerebral hemispheres, which could then be padded and transformed back into native participant space.

A 3D Unet-style architecture^39^ was used, implemented in Keras using Tensorflow 1.15 (see Suppl Fig 2). Four encoding levels were used, each containing two batch normalisation and convolutional layers, and one dropout layer, with dropout set to 50%. Pooling was performed using strided convolutions of the first convolutional layer in each encoding layer to allow the network to learn the optimal means by which to select features. Four decoding layers each consisted of two batch normalisation and convolutional layers with a custom written tri-linear interpolation resizing layer. As with traditional Unet architectures, input into each decoding layer included the linearly upsampled output of the previous decoding layer, concatenated with the output of the corresponding encoding layer. ReLu activation functions were used for all convolutional layers, excepting the final layer, which was softmax.

Training (n=98), testing (n=20), and validation (n=40) datasets were derived from the participants who were used to generate probabilistic images of the temporal lobe and V1/V2 (see main text). Ground truth images were binarised trackmaps generated by seeding from the optic chiasm by the means described earlier, using the Dice coefficient for the loss function. To reduce the probability of over-fitting input images were augmented on the fly using a combination of smoothing, Gaussian noise addition, channel drop-out, left-right flipping and semi-random non-linear geometric distortions. Once trained, the mean Dice coefficients for the validation and test datasets were each ∼0.95. Of the 500 HCP-M90 participants for whom the optic radiation was delineated, 54 also appeared in this training set. No participants included in other datasets contributed to the training, test, or validation datasets.

**Suppl Fig 2.**
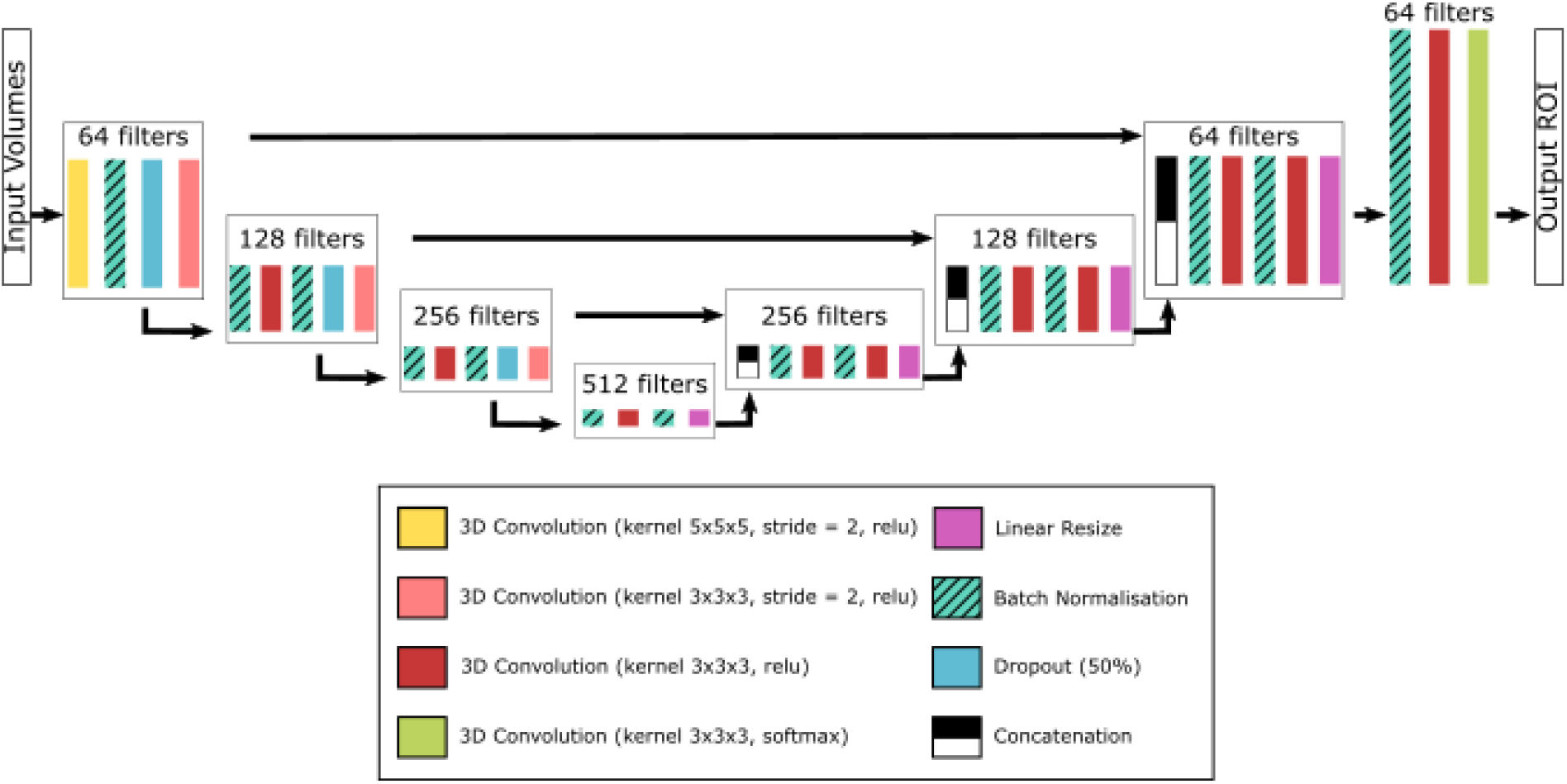
Neural Network Architecture.

**Suppl Fig 3.**
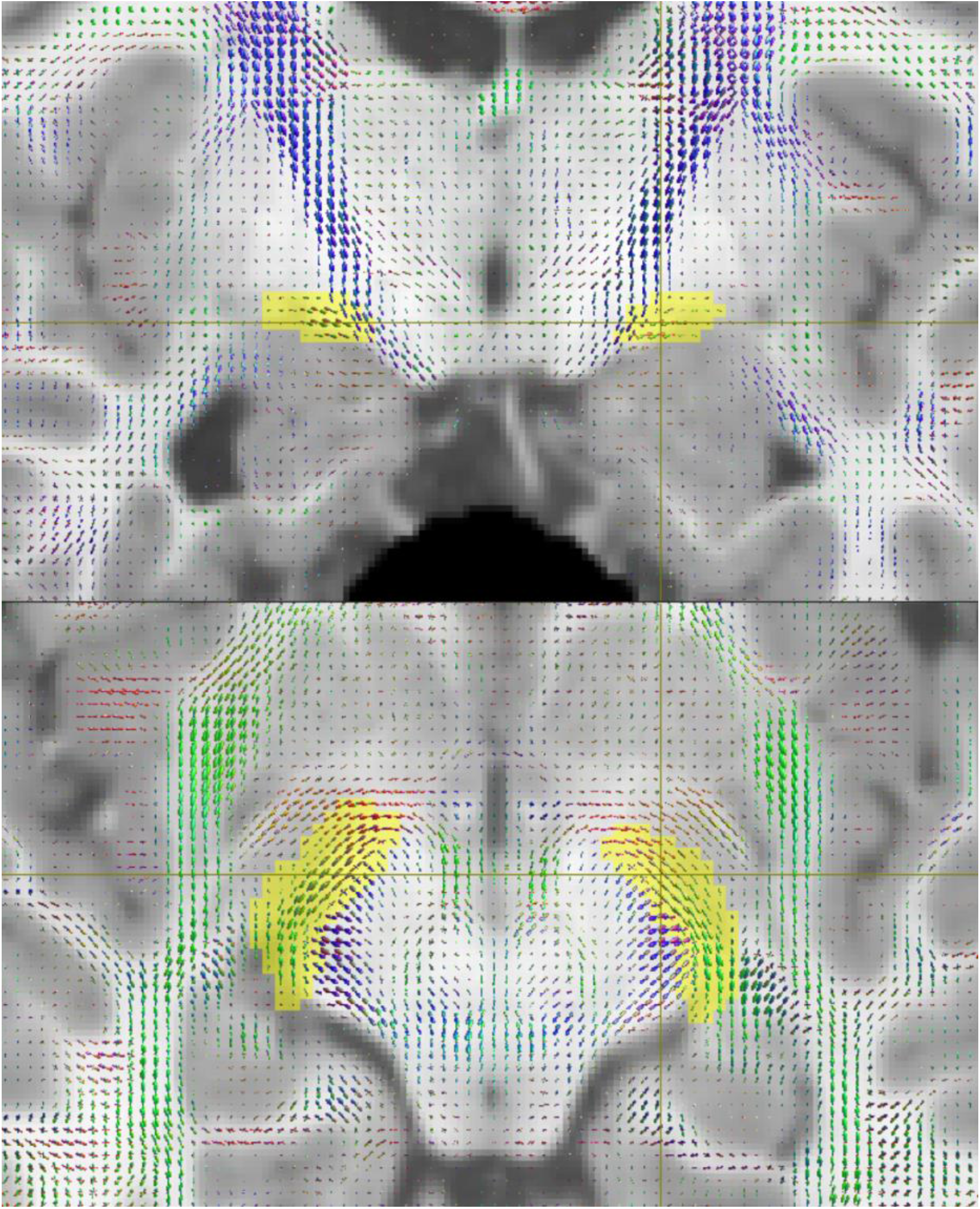
Example of seeding areas (yellow) for the optic radiation at the level of the brainstem, generated using a convolutional neural network. The background image is a T1 MPRAGE (grey) with coloured fibre orientation distributions overlaid, both of which were fed into the network.

**Suppl Fig 4.**
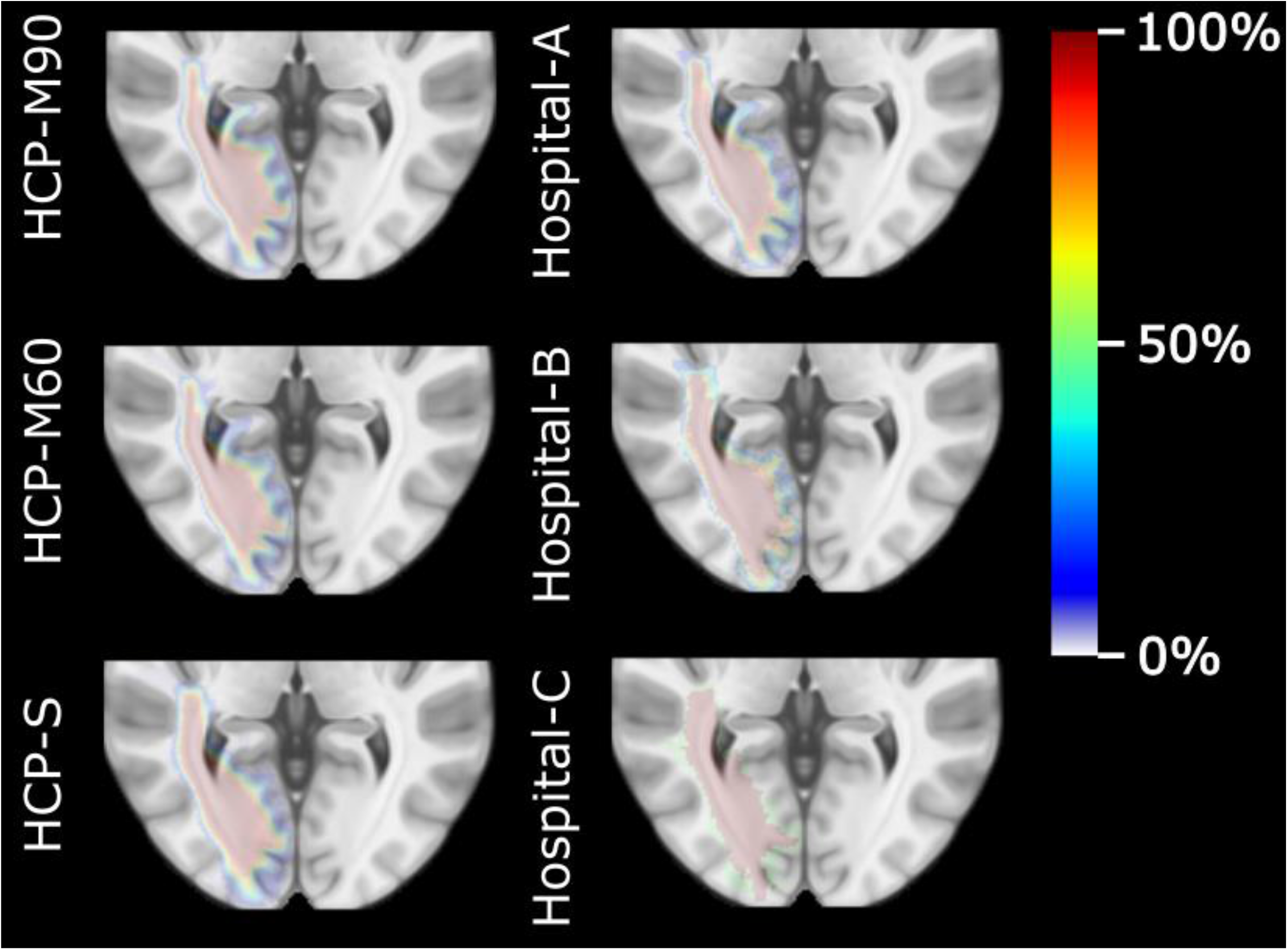
Population tractography ‘heat map’ (displayed as a percentage of participants) overlaid on an MPRAGE MNI template. The border of the middle and posterior optic radiation is visible as a dark line on the MPRAGE image. Tractography using CONSULT typically bordered close to this line, with tighter delineations for acquisitions with higher spatial and angular resolution.

## Notes

### Competing Interest Statement

The authors have declared no competing interest.

### Author Declarations

Ethics was obtained by the appropriate bodies for: 1. Metro North, Queensland, Australia 2. Hospital Clinic, Barcelona, Spain 3. CSIRO, Australia

